# Critically ill patients with cirrhosis outside the unified ACLF definition: a two-database cohort study

**DOI:** 10.64898/2026.11.14.26368094

**Authors:** Zhiqiang Hu, Jihui Ye

**Author notes:** Corresponding author: Jihui Ye, Department of Critical Care Medicine, The First Affiliated Hospital of Ningbo University, Ningbo, China. First author. Full mailing address: 59 Liuting Street, Haishu District, Ningbo 315010, Zhejiang, China.

## Abstract

**Background:** The 2026 multisociety definition of acute-on-chronic liver failure (ACLF) requires bilirubin ≥5 mg/dL and international normalized ratio ≥1.5 before grading extrahepatic organs. In intensive care unit (ICU) patients with cirrhosis, extrahepatic organ failure without liver failure is common. The proportion excluded and their outcomes are unknown.

**Methods:** We applied unified grading and the European Association for the Study of the Liver–Chronic Liver Failure (EASL-CLIF) criteria around ICU admission to adults with coded cirrhosis in the Medical Information Mart for Intensive Care IV (MIMIC-IV), with replication in the eICU Collaborative Research Database (eICU-CRD). In patients positive by EASL-CLIF alone, we estimated cumulative incidence of a documented liver-failure threshold by ICU day 7, with ICU discharge, death and, in MIMIC-IV, transplantation as competing events.

**Results:** Of 2,972 MIMIC-IV patients, 33.8% met unified and 65.2% met EASL-CLIF criteria. Of EASL-CLIF-positive patients, 1,086 (56.0%) were assigned unified grade 0. Of these, 54.7% received invasive ventilation, 48.7% received vasopressors, and 32.6% (95% confidence interval 29.9–35.4%) died within 28 days, compared with 11.8% of patients negative by both. Unified grade 0 contained 46.7% of all 28-day deaths. By day 7, the threshold had been documented in 12.3% (10.5–14.4%) of the 1,086, and 645 (59.4%) had left the ICU alive first. In eICU-CRD, 62.4% of EASL-CLIF-positive patients were assigned grade 0, and the day-7 incidence was 9.6% (6.7–12.5%).

**Conclusion:** About half of patients with ACLF by EASL-CLIF criteria were assigned unified grade 0 in the ICU. One in three died within 28 days, and few reached the liver-failure threshold during the first ICU week. Critical care studies adopting the unified definition should report this group separately.

## Introduction

Acute-on-chronic liver failure (ACLF) describes an abrupt deterioration of chronic liver disease with organ failure and a high short-term risk of death,^1^ but for more than a decade it was defined within three main frameworks. The Asian Pacific Association for the Study of the Liver (APASL) definition centered on jaundice and coagulopathy after an acute hepatic insult.^2^ The European Association for the Study of the Liver–Chronic Liver Failure (EASL-CLIF) definition graded hepatic and extrahepatic organ failures in acutely decompensated cirrhosis and did not require liver failure.^3^ The North American Consortium for the Study of End-Stage Liver Disease (NACSELD) definition required two extrahepatic organ failures.^4,5^ The same patient could have ACLF under one definition and not another.^6,7^

In October 2026, five liver societies agreed on a single definition.^8^ Liver failure, a bilirubin ≥5 mg/dL together with an international normalized ratio (INR) ≥1.5, is now the primary requisite condition; severity is graded by extrahepatic organ dysfunction or failure, and clinical types A and B are distinguished by the history of decompensation.^8-11^ The definition makes ACLF a syndrome of liver failure, and its authors have argued that extrahepatic failure alone, however severe, does not constitute it.^12^

That choice matters most in the intensive care unit (ICU). Patients with cirrhosis admitted to the ICU frequently require mechanical ventilation, vasopressor support or renal replacement therapy,^13,14^ and extrahepatic organ failure may develop without liver failure. Under EASL-CLIF criteria many of them have ACLF; under the unified definition they may not, and the consensus gives this group no separate name or care pathway.^8^ It does recommend that critically ill patients be reassessed on days 3–7,^8^ and the APASL position paper on critical care advises repeated prognostic assessment on days 0, 3, 7 and 14.^15^ It is not known how many of these patients go on to meet the liver criteria during that period. If most do, the unified definition mainly delays the diagnosis. If few do, a large high-risk group remains outside ACLF throughout the early ICU course.

We applied the unified grade and the EASL-CLIF criteria to the same adults with cirrhosis around ICU admission in two intensive care databases. We asked how many patients were classified differently, how often they died, and how many of those positive by EASL-CLIF alone reached the liver-failure threshold during the first week in the ICU.

## Methods

### Data sources and patients

The primary cohort came from the Medical Information Mart for Intensive Care IV (MIMIC-IV) v3.1, which holds deidentified records of patients admitted to Beth Israel Deaconess Medical Center, Boston, from 2008 to 2022.^16,17^ We included adults with a prespecified diagnosis code for cirrhosis, informed by a published code set,^18^ at their first ICU stay, provided that bilirubin and INR were both measured between 24 hours before ICU admission and 24 hours after it or ICU discharge, whichever came first. We excluded patients with a coded previous liver transplant and those with an index-admission transplant on or before the calendar date of ICU admission. Replication used the eICU Collaborative Research Database (eICU-CRD) v2.0, which covers ICU stays in 208 US hospitals in 2014–2015,^19,20^ with the same laboratory window and grade thresholds and database-specific mappings; transplant exclusions were applied only in MIMIC-IV. The study follows the STROBE and RECORD statements.^21,22^

### Classifications

All classifications were derived from routinely recorded laboratory values, vital signs and treatments. Each organ system was assessed using the worst value, or any use of the relevant organ support, in the observation window; urine output, Glasgow Coma Scale scores and invasive pressures were charted only after ICU admission. Acute kidney injury was staged by KDIGO criteria using urine output and creatinine compared with the lowest value in the preceding 48 hours or 7 days, including results from earlier hospital encounters. We call our implementation of the consensus grading the unified grade.^8^ Patients who met both hepatic thresholds were assigned grade 1 if they had extrahepatic organ dysfunction without failure, grade 2 with one extrahepatic organ failure and grade 3 with two or more; all others were assigned grade 0. Kidney dysfunction was defined as Kidney Disease: Improving Global Outcomes (KDIGO) acute kidney injury stage 1 and kidney failure as stage 2 or higher or new renal replacement therapy (RRT).^23^ Circulatory dysfunction was a mean arterial pressure below 65 mm Hg without vasopressors, and circulatory failure any vasopressor use. Respiratory dysfunction was a PaO_2_/FiO_2_ ratio of more than 225 to 300 (or an SpO_2_/FiO_2_ ratio of more than 244 to 333 when PaO_2_ was unavailable), and respiratory failure a lower ratio or invasive mechanical ventilation. West Haven grade is not recorded, so cerebral failure was approximated by a Glasgow Coma Scale (GCS) score ≤13, as in earlier ICU database studies,^24-26^ and cerebral dysfunction by a score of 14. Types A and B could not be assigned, because the history and timing of decompensation are not available.

EASL-CLIF grade, the CLIF Consortium Organ Failure (CLIF-C OF) score, NACSELD ACLF, MELD 3.0 and the Sequential Organ Failure Assessment (SOFA) score were calculated from the same data.^3,5,27-29^ Because the EASL-CLIF definition applies to acute decompensation, which cannot be confirmed from these records, we refer to the EASL-CLIF criteria as applied to ICU patients with coded cirrhosis. Unmeasured nonhepatic components were scored as normal. Suspected infection was defined by microbiological culture sampling paired with an antimicrobial prescription within 24 hours of ICU admission, a definition that also captures prophylactic antibiotics; infection and decompensating events (variceal bleeding, hepatic encephalopathy, ascites, spontaneous bacterial peritonitis and hepatorenal syndrome) were also taken from index-hospitalization diagnosis codes.

Variable mappings and code lists are given in Supplemental Methods.

### Course over the first week

Patients positive by EASL-CLIF alone were followed from ICU admission to 168 hours. Days were counted in 24-hour blocks from admission, with day 2 covering 24 to less than 48 hours and day 7 covering 144 to less than 168 hours. From day 2 onwards, a day with both bilirubin and INR measured before ICU exit, death or transplantation (where ascertainable) was evaluable, and the liver-failure threshold was observed on the first day on which both values met it. We estimated the cumulative incidence of a first observed threshold with the Aalen–

Johansen estimator, treating ICU discharge alive, death in the ICU and liver transplantation as competing events.^30^ For each day we also recorded whether patients still in the ICU had paired tests below the threshold or no paired tests. In patients who reached the threshold, extrahepatic organs were assessed on the same day to determine whether unified ACLF criteria were met. Among patients alive at day 7, 28-day mortality was described by whether the threshold had been observed. In eICU-CRD, the timing of transplantation could not be established, and the few patients whose status at ICU discharge was not recorded were censored. To describe what followed ICU discharge, we recorded where deaths by day 28 occurred and discharges to hospice.

### Outcomes and statistical analysis

The primary outcome was death from any cause within 28 days of ICU admission. Secondary outcomes were death within 90 days and during the index hospitalization; in eICU-CRD, in-hospital death was the only outcome. Deaths in MIMIC-IV were ascertained from hospital records and linked state death records.^17^

We report the cross-classification of unified and EASL-CLIF grades and four agreement groups: negative by both, positive by EASL-CLIF alone, positive by the unified grade alone and positive by both. Proportions are given with Wilson 95% confidence intervals (CIs).^31^ Discrimination for 28-day mortality was measured by the area under the receiver operating characteristic curve (AUC), and the definitions were compared both as binary criteria (ACLF present or absent) and as four-level grades, with 2,000 paired bootstrap resamples.^32^ The continuous CLIF-C OF score is reported as a prognostic reference. We also cross-classified the unified grade against NACSELD criteria.

Sensitivity analyses required impaired oxygenation for respiratory failure, excluded patients intubated or sedated at admission, omitted the cerebral domain, varied the handling of transplantation, excluded elective admissions and restricted the windows to the first 24 or 48 hours. A post hoc analysis restricted the cohort to patients with a diagnosis code for ascites, hepatic encephalopathy, esophageal variceal bleeding, hepatorenal syndrome, spontaneous bacterial peritonitis or jaundice during the index hospitalization. The first-week analyses, prompted by the consensus recommendation to reassess critically ill patients on days 3–7, and three further sensitivity analyses were post hoc: GCS 14 was no longer counted as cerebral dysfunction; bilirubin and INR were required to be sampled within 12 hours of each other; and patients with two or more extrahepatic dysfunctions without failure were assigned grade 0 rather than grade 1, a situation the consensus does not address.

Analyses used Python 3.14.6 with DuckDB, pandas, NumPy, SciPy and statsmodels. Codex (GPT-6.1 Sol, OpenAI) and Claude (Opus 5.5, Anthropic) assisted with English-language editing and grammatical revision of this manuscript. The authors made all scientific decisions, approved each stage of the analysis, verified the results and take full responsibility for the content.

### Ethics

The Beth Israel Deaconess Medical Center Institutional Review Board reviewed the collection of patient information and creation of MIMIC-IV, waived informed consent and approved data sharing.^16,17^ eICU-CRD was deidentified under the HIPAA Safe Harbor provision, and Privacert certified the re-identification risk as meeting that standard (HIPAA Certification No. 1031219-2).^19,20^ This secondary analysis used deidentified data obtained through credentialed access under each database’s data use agreement and met the conditions for exemption from ethics review under Article 32(2) of China’s Ethical Review Measures for Life Sciences and Medical Research Involving Human Beings.^33^ No study-specific approval or exemption letter was issued. The study followed the Declaration of Helsinki.

## Results

### Patients

Of 3,806 adults with coded cirrhosis at their first ICU stay in MIMIC-IV, 3,232 had bilirubin and INR measured in the observation window, and 2,972 remained after exclusion of 260 with a coded previous transplant or a transplant on or before the ICU admission date (Supplemental Figure S1). The median age was 59 years, the median MELD 3.0 score 25, the median bilirubin 3.2 mg/dL and the median SOFA score 7 (Table 1). A decompensation-related diagnosis was coded during the hospitalization in 1,883 patients (63.4%) and infection or sepsis in 1,730 (58.2%). By day 28, 980 patients (33.0%) had died.

**Table 1.** Characteristics of the cohort at entry to intensive care.

| **Characteristic** | **Overall<br>(n=2,972)** | **Grade 0<br>(n=1,968)** | **Grade 1<br>(n=202)** | **Grade 2<br>(n=290)** | **Grade 3<br>(n=512)** |
|---|---|---|---|---|---|
| Age, years | 59 (51, 67) | 61 (54, 70) | 54 (46, 61) | 55 (49, 61) | 55 (47, 62) |
| Female | 1,078 (36.3%) | 688 (35.0%) | 79 (39.1%) | 109 (37.6%) | 202 (39.5%) |
| Alcohol-related cirrhosis | 1,678 (56.5%) | 966 (49.1%) | 136 (67.3%) | 213 (73.4%) | 363 (70.9%) |
| Suspected infection around ICU admission | 2,209 (74.3%) | 1,416 (72.0%) | 141 (69.8%) | 218 (75.2%) | 434 (84.8%) |
| Infection or sepsis coded during index hospitalization | 1,730 (58.2%) | 999 (50.8%) | 127 (62.9%) | 200 (69.0%) | 404 (78.9%) |
| Intubated at ICU entry | 664 (22.3%) | 438 (22.3%) | <10 | 40 (13.8%) | 182 (35.5%) |
| Bilirubin, mg/dL | 3.2 (1.4, 8.4) | 1.8 (1, 3.2) | 10.3 (6.83, 17.8) | 11.85 (7.03, 22.18) | 13.3 (7.9, 24.2) |
| INR | 1.8 (1.4, 2.4) | 1.6 (1.3, 1.9) | 2.2 (1.9, 2.8) | 2.3 (1.9, 3) | 2.5 (2.1, 3.3) |
| Creatinine, mg/dL | 1.4 (0.9, 2.5) | 1.2 (0.8, 2.1) | 1.2 (0.9, 2) | 1.6 (1, 2.77) | 2.4 (1.4, 4.2) |
| MELD 3.0 | 25 (17, 33) | 20 (15, 26) | 31 (26.25, 36) | 33 (28, 38.75) | 37 (31, 40) |
| SOFA | 7 (5, 11) | 6 (4, 8) | 7 (6, 8) | 9 (7, 11) | 13 (11, 15) |
| Lactate, mmol/L | 2.7 (1.8, 4.7) | 2.4 (1.7, 4.1) | 2.45 (1.8, 4) | 2.8 (1.9, 4.2) | 4 (2.4, 7.5) |

Values are median (first quartile, third quartile) or number (percentage). Grade refers to unified grading. Counts below 10 are shown as <10 and percentages are omitted.
Suspected infection: microbiological culture sampling paired with an antimicrobial prescription within 24 hours of ICU admission; this definition also captures prophylactic antibiotics. Diagnosis codes cover the index hospitalization.
ICU, intensive care unit. INR, international normalized ratio. MELD, Model for End-Stage Liver Disease. SOFA, Sequential Organ Failure Assessment.

### Reclassification and mortality

ACLF criteria were met by 1,004 patients (33.8%) under the unified grade and by 1,938 (65.2%) under EASL-CLIF; NACSELD criteria were met by 29.9% (Supplemental Table S2). The two classifications disagreed in 1,238 patients, almost always in one direction: 1,086 were positive by EASL-CLIF alone and 152 by the unified grade alone (Figure 1A). All 152 met both hepatic thresholds. In 78 of them, none of the recorded extrahepatic findings reached EASL-CLIF organ failure; the most common pattern was isolated hypotension without vasopressors, which EASL-CLIF does not count. The other 74 had a single non-renal organ failure without the kidney dysfunction or encephalopathy that EASL-CLIF grade 1 requires (Supplemental Table S4). The 1,086 made up 56.0% of all EASL-CLIF-positive patients and included 305 with EASL-CLIF grade 3 (Supplemental Figure S2). Unified grade 0 also included 504 patients who met NACSELD criteria, of whom 37.5% died within 28 days (Supplemental Table S2).

**Figure 1.**
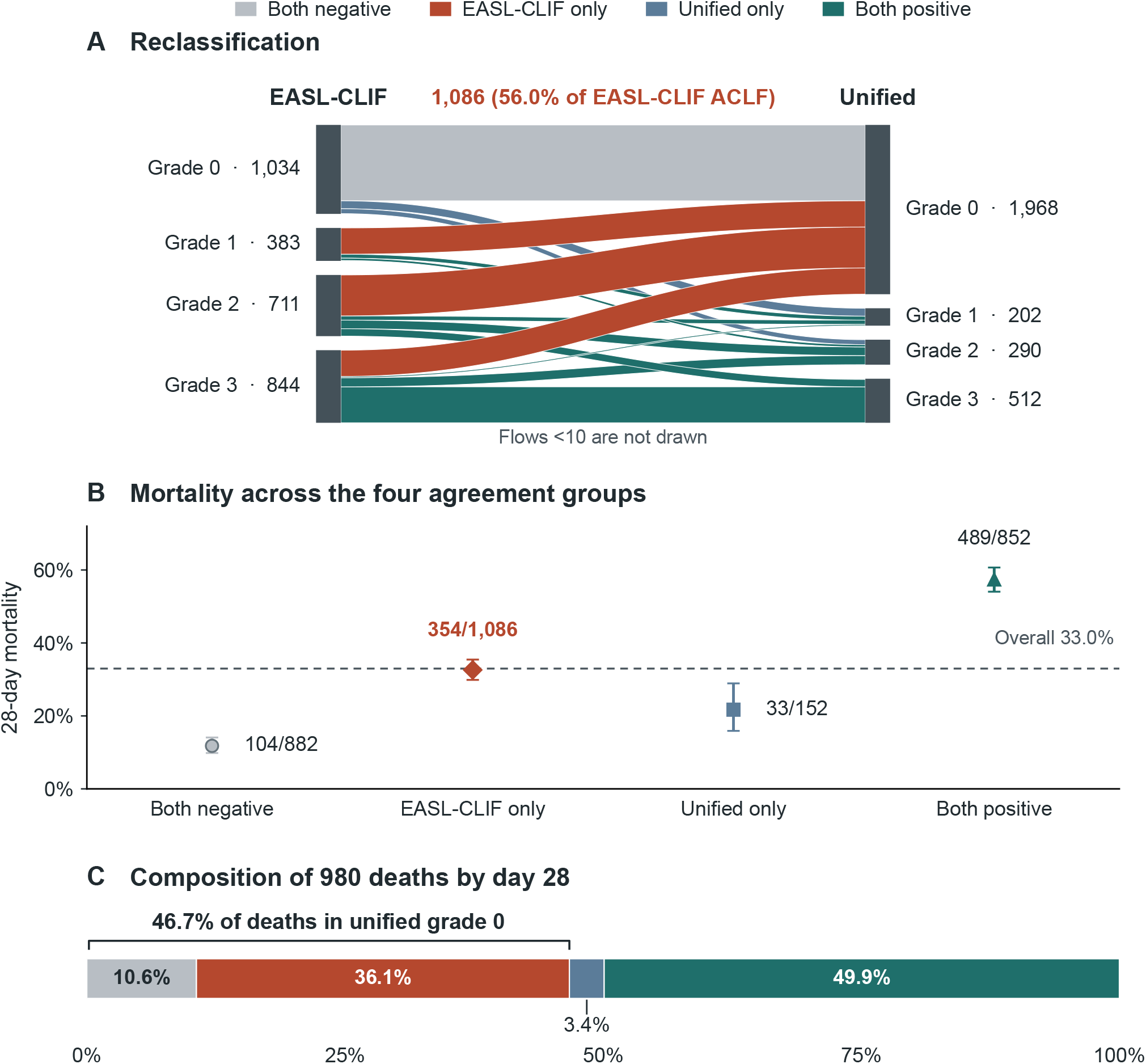
Reclassification and 28-day mortality in MIMIC-IV. (A) Alluvial plot connecting EASL-CLIF grades with unified grades in the 2,972 patients. Node totals include every patient; ribbons are drawn for flows of at least 10 patients. Colors indicate the four agreement groups. (B) 28-day mortality by agreement group, with error bars showing Wilson 95% CIs; labels give deaths over group size, and the dashed line marks overall mortality (33.0%). (C) Distribution of the 980 deaths by agreement group; the bracket spans the deaths at unified grade 0. The full cross-classification is given in Supplemental Figure S2.

Patients positive by EASL-CLIF alone had little biochemical liver failure but substantial organ support (Table 2). Their median bilirubin was 1.9 mg/dL, and 1,067 of them fell below one or both hepatic thresholds; the other 19 met both thresholds but had no recorded extrahepatic dysfunction, and in 13 of them EASL-CLIF ACLF rested on concurrent liver and coagulation failure (Supplemental Table S4). Their organ failures were mainly extrahepatic: more than half received invasive ventilation, nearly half received vasopressors and about half had EASL-CLIF kidney failure. Ascites was coded during the hospitalization in 46.6% and infection or sepsis in 62.2%. Their 28-day mortality was 32.6% (95% CI 29.9–35.4%), nearly three times that of patients negative by both classifications (11.8%) and higher than that of patients positive by the unified grade alone (21.7%) (Figure 1B).

**Table 2.** Clinical characteristics and mortality by agreement group.

| **Characteristic** | **Both negative<br>(n = 882)** | **EASL-CLIF only<br>(n = 1,086)** | **Unified only<br>(n = 152)** | **Both positive<br>(n = 852)** |
|---|---|---|---|---|
| Bilirubin, mg/dL | 1.75 (1, 3) | 1.9 (1, 3.3) | 7.3 (5.88, 9.9) | 13.8 (8.1, 23.73) |
| INR | 1.5 (1.3, 1.7) | 1.7 (1.4, 2.18) | 2 (1.77, 2.2) | 2.5 (2.1, 3.3) |
| MELD 3.0 | 16 (12, 20) | 24 (18, 30) | 25 (23, 28) | 36 (31, 40) |
| SOFA | 4 (3, 6) | 8 (6, 10) | 7 (5.75, 7.25) | 11 (9, 14) |
| Lactate, mmol/L | 2.1 (1.6, 3) | 2.8 (1.8, 4.9) | 2.6 (1.9, 3.58) | 3.5 (2.2, 6.2) |
| Suspected infection around ICU admission | 564 (63.9%) | 852 (78.5%) | 107 (70.4%) | 686 (80.5%) |
| Ascites coded | 304 (34.5%) | 506 (46.6%) | 94 (61.8%) | 622 (73.0%) |
| Spontaneous bacterial peritonitis coded | 38 (4.3%) | 100 (9.2%) | 13 (8.6%) | 165 (19.4%) |
| Hepatorenal syndrome coded | 18 (2.0%) | 123 (11.3%) | 16 (10.5%) | 298 (35.0%) |
| Esophageal variceal bleeding coded | 142 (16.1%) | 111 (10.2%) | 18 (11.8%) | 122 (14.3%) |
| Hepatic encephalopathy coded | 64 (7.3%) | 109 (10.0%) | 21 (13.8%) | 147 (17.3%) |
| Fresh frozen plasma use | 69 (7.8%) | 205 (18.9%) | 37 (24.3%) | 308 (36.2%) |
| Organ support, n (%) |  |  |  |  |
| Vasopressor use | 35 (4.0%) | 529 (48.7%) | <10 | 387 (45.4%) |
| Invasive ventilation | 160 (18.1%) | 594 (54.7%) | 28 (18.4%) | 403 (47.3%) |
| RRT | <10 | 72 (6.6%) | <10 | 90 (10.6%) |
| Liver failure | 11 (1.2%) | 28 (2.6%) | 14 (9.2%) | 491 (57.6%) |
| Kidney failure | <10 | 570 (52.5%) | <10 | 488 (57.3%) |
| Brain failure | 92 (10.4%) | 277 (25.5%) | 15 (9.9%) | 298 (35.0%) |
| Coagulation failure | 18 (2.0%) | 190 (17.5%) | <10 | 463 (54.3%) |
| Circulatory failure | 35 (4.0%) | 529 (48.7%) | <10 | 387 (45.4%) |
| Respiratory failure | 177 (20.1%) | 671 (61.8%) | 33 (21.7%) | 473 (55.5%) |
| 28-day mortality | 104/882 (11.8%)<br>(9.8%–14.1%) | 354/1,086 (32.6%)<br>(29.9%–35.4%) | 33/152 (21.7%)<br>(15.9%–28.9%) | 489/852 (57.4%)<br>(54.0%–60.7%) |
| 90-day mortality | 168/882 (19.0%)<br>(16.6%–21.8%) | 466/1,086 (42.9%)<br>(40.0%–45.9%) | 54/152 (35.5%)<br>(28.4%–43.4%) | 567/852 (66.5%)<br>(63.3%–69.6%) |
| In-hospital mortality | 72/882 (8.2%)<br>(6.5%–10.2%) | 296/1,086 (27.3%)<br>(24.7%–30.0%) | 28/152 (18.4%)<br>(13.1%–25.3%) | 453/852 (53.2%)<br>(49.8%–56.5%) |

Laboratory and severity measures are median (first quartile, third quartile). Organ support and EASL-CLIF organ failures are n (%). Organ domains can overlap. Positive classification is grade 1 or higher.
Lactate summaries use measured values. Available measurements by column were 517, 925, 98, and 741.
Suspected infection: microbiological culture sampling paired with an antimicrobial prescription within 24 hours of ICU admission; this definition also captures prophylactic antibiotics. Decompensation-related codes record diagnoses during the index hospitalization. Plasma use is a recorded administration or infusion within the window from 24 hours before to less than 24 hours after ICU admission.
Mortality cells show deaths divided by the outcome denominator (percentage) and the Wilson 95% confidence interval on the line below. Low counts are shown as <10 with the corresponding percentages and intervals omitted.
EASL-CLIF, European Association for the Study of the Liver–Chronic Liver Failure. INR, international normalized ratio. MELD, Model for End-Stage Liver Disease. RRT, renal replacement therapy. SOFA, Sequential Organ Failure Assessment.

As a result, unified grade 0 contained 458 of the 980 deaths by day 28 (46.7%), 354 of them among patients positive by EASL-CLIF alone (Figure 1C). Above grade 0, mortality rose steeply, from 29.2% at grade 1 to 65.6% at grade 3, but grade 0 held two thirds of the cohort and combined two groups with very different risks (Supplemental Figure S3A).

### Course over the first week

In patients positive by EASL-CLIF alone, the liver-failure threshold had been documented in 6.3% by day 2 and in 12.3% (95% CI 10.5–14.4%) by day 7 (Figure 2C). Nearly all of these patients (126 of 134) had extrahepatic dysfunction or failure recorded on the same day and thus met our operational criteria for unified grade 1 or higher. More often, patients left the ICU first: 645 were discharged alive before the threshold was documented (Figure 2A). Of these 645 patients, 112 (17.4%) died by day 28: 51 later in the same hospitalization, 55 after hospital discharge and fewer than 10 at a time that could not be assigned. Forty-one were discharged from hospital to hospice. Of the 186 patients still in the ICU at 168 hours without a documented threshold, 105 had paired tests below it on day 7. eICU-CRD showed the same pattern, with a day-7 cumulative incidence of 9.6% (6.7–12.5%) among its 343 patients positive by EASL-CLIF alone (Figure 2B, C).

**Figure 2.**
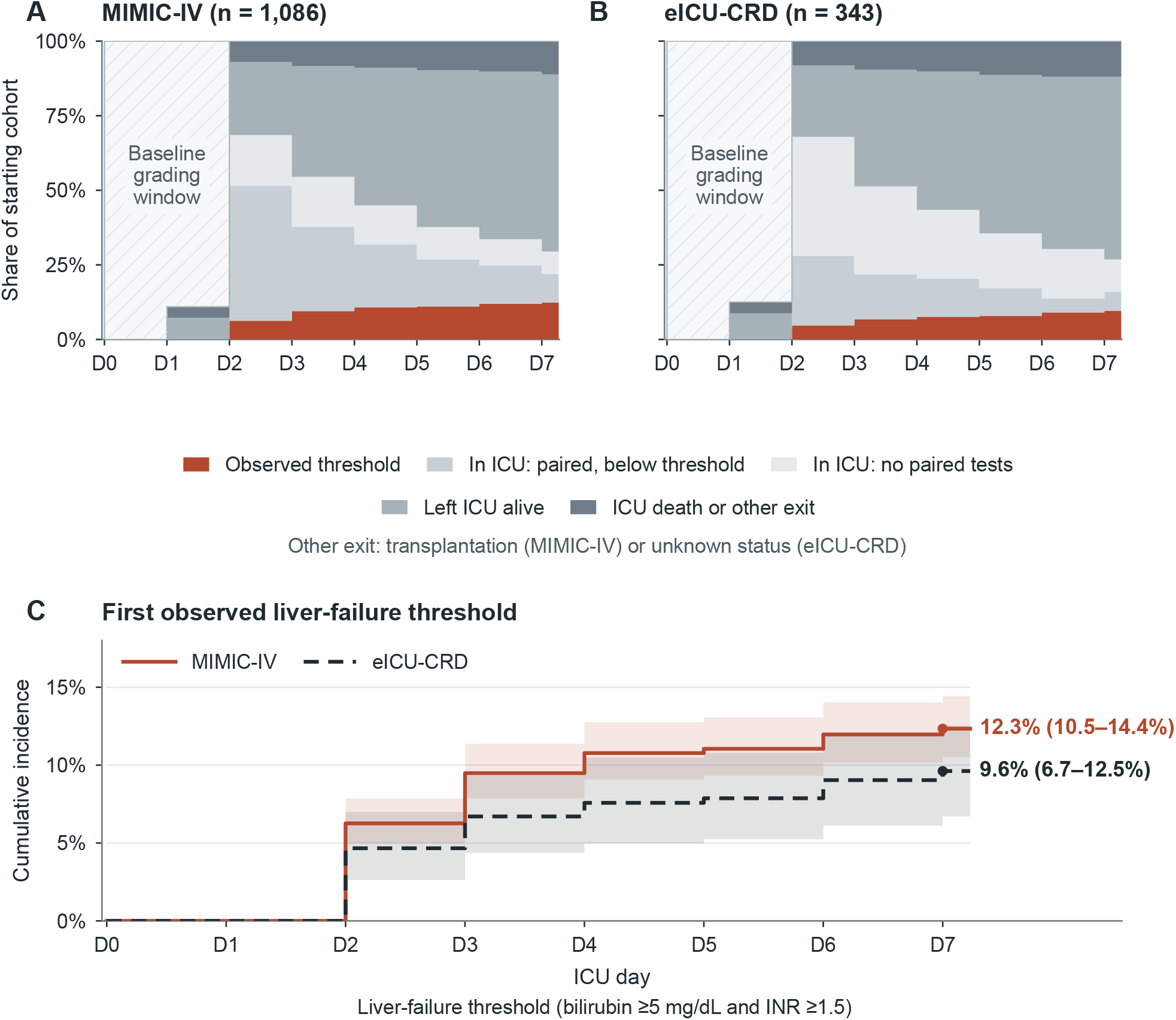
Course over the first ICU week in patients positive by EASL-CLIF alone. (A, B) Daily state of patients in MIMIC-IV (n = 1,086) and eICU-CRD (n = 343): liver-failure threshold observed, paired tests below the threshold, no paired tests, discharged from the ICU alive, and ICU death or other exit (transplantation in MIMIC-IV or unknown status in eICU-CRD). The hatched band marks the baseline grading window. (C) Aalen– Johansen cumulative incidence of a first observed liver-failure threshold. Live ICU discharge, ICU death and, in MIMIC-IV, liver transplantation were competing events. ICU discharges of unknown status in eICU-CRD were right-censored at discharge. Shaded bands show pointwise 95% CIs (Wilson in MIMIC-IV and 2,000 patient-level percentile bootstrap resamples in eICU-CRD).

Among patients alive at day 7, 28-day mortality was 38.7% in those in whom the threshold had been documented and 19.1% in those evaluable without it (Supplemental Table S3).

### Discrimination and sensitivity analyses

As binary criteria, the unified and EASL-CLIF classifications had AUCs for 28-day mortality of 0.645 and 0.655 (difference −0.010; 95% CI −0.030 to 0.011). As four-level grades, the unified grade had the lower AUC (0.665 vs 0.736; difference −0.071; −0.090 to −0.050) (Table 3). The continuous CLIF-C OF score, included as a reference, had an AUC of 0.771.

**Table 3.**
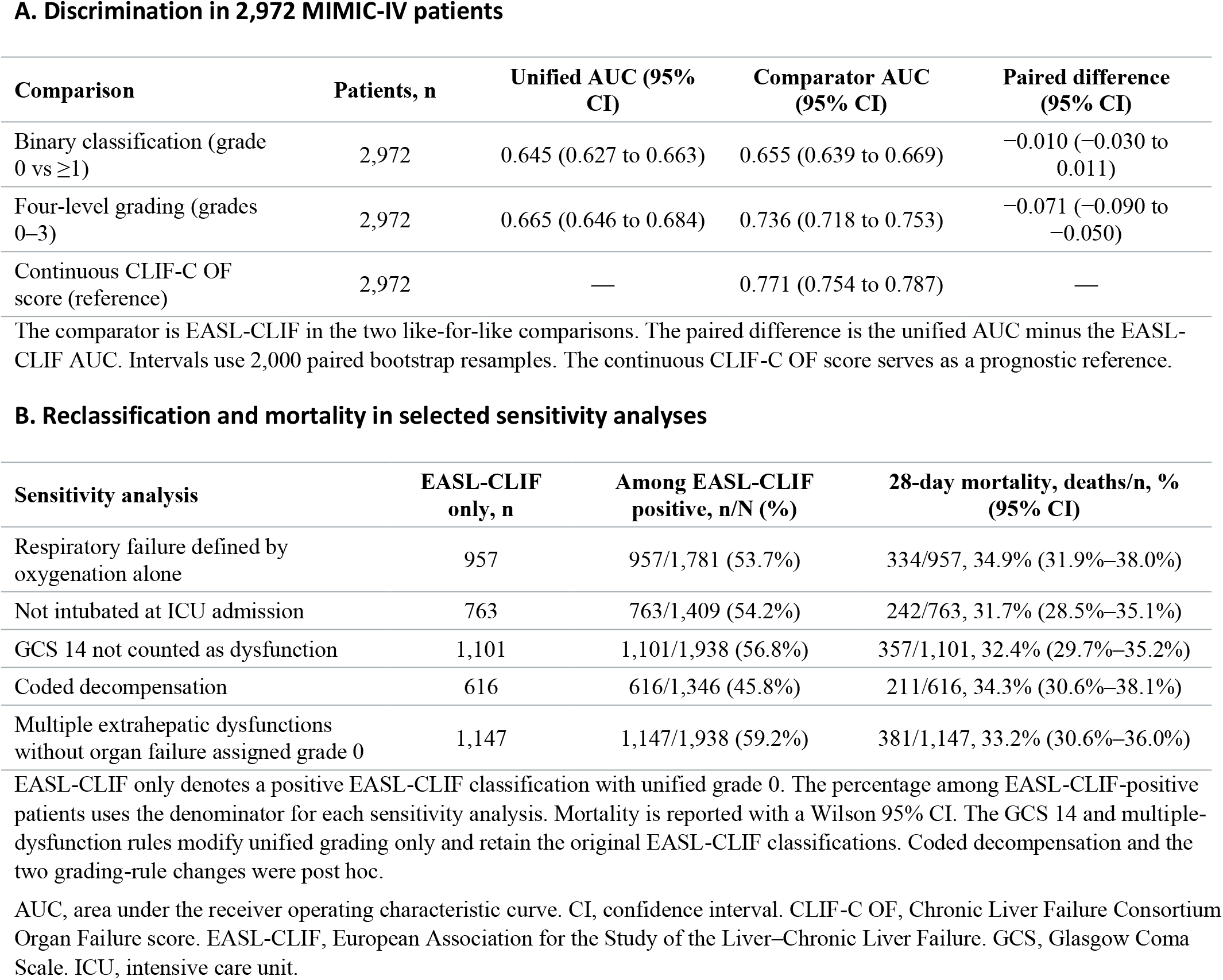
Discrimination for 28-day mortality and sensitivity analyses.

| **A. Discrimination in 2,972 MIMIC-IV patients** |  |  |  |  |
|---|---|---|---|---|
| **Comparison** | **Patients, n** | **Unified AUC (95% CI)** | **Comparator AUC (95% CI)** | **Paired difference (95% CI)** |
| Binary classification (grade 0 vs ≥1) | 2,972 | 0.645 (0.627 to 0.663) | 0.655 (0.639 to 0.669) | −0.010 (−0.030 to 0.011) |
| Four-level grading (grades 0–3) | 2,972 | 0.665 (0.646 to 0.684) | 0.736 (0.718 to 0.753) | −0.071 (−0.090 to −0.050) |
| Continuous CLIF-C OF score (reference) | 2,972 | — | 0.771 (0.754 to 0.787) | — |
| **B. Reclassification and mortality in selected sensitivity analyses** |  |  |  |  |
| **Sensitivity analysis** | **EASL-CLIF only, n** | **Among EASL-CLIF positive, n/N (%)** | **28-day mortality, deaths/n, % (95% CI)** |  |
| Respiratory failure defined by oxygenation alone | 957 | 957/1,781 (53.7%) | 334/957, 34.9% (31.9%–38.0%) |  |
| Not intubated at ICU admission | 763 | 763/1,409 (54.2%) | 242/763, 31.7% (28.5%–35.1%) |  |
| GCS 14 not counted as dysfunction | 1,101 | 1,101/1,938 (56.8%) | 357/1,101, 32.4% (29.7%–35.2%) |  |
| Coded decompensation | 616 | 616/1,346 (45.8%) | 211/616, 34.3% (30.6%–38.1%) |  |
| Multiple extrahepatic dysfunctions without organ failure assigned grade 0 | 1,147 | 1,147/1,938 (59.2%) | 381/1,147, 33.2% (30.6%–36.0%) |  |

The comparator is EASL-CLIF in the two like-for-like comparisons. The paired difference is the unified AUC minus the EASL-CLIF AUC. Intervals use 2,000 paired bootstrap resamples. The continuous CLIF-C OF score serves as a prognostic reference.
EASL-CLIF only denotes a positive EASL-CLIF classification with unified grade 0. The percentage among EASL-CLIF-positive patients uses the denominator for each sensitivity analysis. Mortality is reported with a Wilson 95% CI. The GCS 14 and multiple-dysfunction rules modify unified grading only and retain the original EASL-CLIF classifications. Coded decompensation and the two grading-rule changes were post hoc.
AUC, area under the receiver operating characteristic curve. CI, confidence interval. CLIF-C OF, Chronic Liver Failure Consortium Organ Failure score. EASL-CLIF, European Association for the Study of the Liver–Chronic Liver Failure. GCS, Glasgow Coma Scale. ICU, intensive care unit.

The reclassified group, and its mortality, persisted when respiratory failure required impaired oxygenation, when patients intubated at admission were excluded and when the cerebral criteria were changed (Table 3). In the 1,883 patients with a coded decompensation-related diagnosis, 45.8% of EASL-CLIF-positive patients were assigned unified grade 0, and 34.3% of them died within 28 days. Requiring bilirubin and INR within 12 hours of each other changed the unified grade in fewer than 10 patients, and assigning grade 0 to patients with several extrahepatic dysfunctions but no failure left the results essentially unchanged (Supplemental Table S4).

### Replication in eICU-CRD

In eICU-CRD, 1,008 patients from 139 hospitals were included, and 343 of 550 EASL-CLIF-positive patients (62.4%) were assigned unified grade 0. In-hospital mortality was 28.9% in patients positive by EASL-CLIF alone and 8.1% in those negative by both, and unified grade 0 contained 51.8% of in-hospital deaths (Supplemental Table S5).

## Discussion

In this study of adults with cirrhosis admitted to intensive care, the unified definition identified ACLF in about half as many patients as the EASL-CLIF criteria. The patients who made up the difference were seriously ill: half were ventilated, half received vasopressors, and one in three died within 28 days. Because of them, nearly half of all early deaths occurred at unified grade 0. Few of these patients reached the liver-failure threshold during the first week in the ICU, and the same pattern was seen in a multicenter database.

The reclassification follows from the structure of the new definition. The consensus makes liver failure the requisite condition for ACLF and grades extrahepatic organs only once it is present.^8^ In our cohort, organ failure without liver failure was common: the median bilirubin was 3.2 mg/dL overall and 1.9 mg/dL in patients positive by EASL-CLIF alone. Most had decompensated cirrhosis by the usual clinical markers, with coded ascites in nearly half and coded infection or sepsis in most, but their organ failures were predominantly extrahepatic, above all of the kidney, lungs and circulation. The overlap with the North American definition is also partial, with 504 patients who met NACSELD criteria assigned unified grade 0. The finding did not depend on counting every ventilated patient as having respiratory failure. It persisted when impaired oxygenation was required, when patients intubated at admission were excluded and when the cohort was limited to patients with coded decompensation. Under EASL-CLIF or NACSELD criteria many of these patients would have been diagnosed with ACLF; under the unified definition, which describes ACLF as a syndrome of liver failure, they are critically ill patients with cirrhosis but without ACLF. The change of label is not a change in risk: about one in three died within 28 days.

The first-week data address an obvious objection, namely that these patients are only diagnosed later. Had that been so, a substantial share would have been expected to reach the liver-failure threshold within days, while they were still being tested in the ICU. About one in eight did so by day 7, most were discharged from the ICU first, and the proportion was similar in eICU-CRD. The consensus recommends reassessing critically ill patients on days 3–7,^8^ and in a Brazilian ICU cohort graded on admission and on days 3 and 7, classification under earlier definitions changed during this period.^34^ For patients positive by EASL-CLIF alone, our data suggest that such reassessment will seldom move them into unified ACLF while they remain in the ICU. Their risk also continued after ICU discharge. One in six of those who left the ICU alive died within 28 days, half of them after hospital discharge, which argues for follow-up of this group beyond the ICU stay. The minority who did reach the threshold had higher mortality among those alive at day 7, in keeping with the view that liver failure marks a worse course.^10,12^

How the definitions compared in discrimination depended on how they were analyzed. As binary criteria, they differed by 0.010 in AUC, with a confidence interval spanning zero. As four-level grades, the unified grade ranked risk clearly less well, consistent with a grade 0 that combined patients with 28-day mortality of 11.8% and 32.6%. In the ICU, prognostic assessment of patients at unified grade 0 therefore requires a separate measure of extrahepatic organ failure, such as the CLIF-C OF or SOFA score.

Registry and cohort studies of critically ill patients with cirrhosis have shown high mortality that rises with the number of failing organs,^13,14,35,36^ and database studies have used the earlier frameworks to describe organ failure or compare prognostic scores in this setting.^24,25,37,38^ In broader cirrhosis cohorts, definitions that require a biochemical liver threshold select different patients from those based on organ failure,^6,39,40^ and a conference report applying a pragmatic version of the harmonized criteria to three inpatient cohorts found mortality rising with grade.^41^ To our knowledge, the final grading rules have not been examined in a dedicated ICU population, and patients outside the definition have not been followed through the first week.

For clinicians, unified grade 0 should not be read as reassurance in the ICU. Prognosis in critically ill patients with cirrhosis should continue to rest on the number and severity of organ failures, as current guidance advises,^42^ whatever the unified grade. For research, the reclassified group has practical consequences. If entry to ACLF trials is based on the unified grade, about half of the ICU patients who meet EASL-CLIF criteria will be excluded, and cohort studies adopting the new definition will lose sight of them unless they are reported separately. Whether these patients benefit from management pathways developed for ACLF, including early transplant assessment, cannot be answered from our data and needs to be studied directly.

### Limitations

Some limitations concern classification. Cirrhosis was identified from diagnosis codes, and acute decompensation, its timing and the history needed to assign types A and B could not be established; restriction to patients with coded decompensation lowered the reclassified share from 56.0% to 45.8% with similar mortality. West Haven grade was approximated by the GCS, which can register sedation or non-hepatic coma; in patients positive by EASL-CLIF alone, cerebral failure by this rule was recorded in 25.5%, whereas hepatic encephalopathy was coded in 10.0%, and the indications for ventilation and vasopressors could not be adjudicated, although the results held when the cerebral criteria were changed and when respiratory failure required impaired oxygenation.

Others concern the data and their timing. Grades used the worst available values in a window of up to 48 hours around admission, and unmeasured components were scored as normal. Plasma was given around admission to 18.9% of patients positive by EASL-CLIF alone, which may have lowered the INR below 1.5 in some, and the timing of coded infections and decompensating events could not be established. Transitions to comfort care are poorly captured in structured records; some ICU discharges and later ward deaths may reflect such transitions, after which laboratory testing usually stops. The liver-failure threshold could be documented only while patients were tested in the ICU, so the cumulative incidence describes what was observed there, not the course after discharge. The primary cohort came from one center; eICU-CRD provided only in-hospital outcomes and no transplant timing, and deaths after discharge may be incompletely captured in MIMIC-IV. The first-week analyses and three sensitivity analyses were defined after the initial results were known.

## Conclusion

In critically ill patients with cirrhosis, the unified ACLF definition assigned grade 0 to about half of those meeting EASL-CLIF criteria. These patients had high short-term mortality and seldom reached the liver-failure threshold during the first week in the ICU. Critical care studies that adopt the unified definition should report them separately.

## Abbreviations

ACLF: acute-on-chronic liver failure
APASL: Asian Pacific Association for the Study of the Liver
AUC: area under the receiver operating characteristic curve
CI: confidence interval
CLIF-C OF: CLIF Consortium Organ Failure
EASL-CLIF: European Association for the Study of the Liver–Chronic Liver Failure
eICU-CRD: eICU Collaborative Research Database
EHOD: extrahepatic organ dysfunction
EHOF: extrahepatic organ failure
GCS: Glasgow Coma Scale
HIPAA: Health Insurance Portability and Accountability Act
ICU: intensive care unit
INR: international normalized ratio
MELD: Model for End-Stage Liver Disease
MIMIC-IV: Medical Information Mart for Intensive Care IV
NACSELD: North American Consortium for the Study of End-Stage Liver Disease
RECORD: REporting of studies Conducted using Observational Routinely collected health Data
RRT: renal replacement therapy
SDC: Supplemental Digital Content
SOFA: Sequential Organ Failure Assessment
STROBE: Strengthening the Reporting of Observational Studies in Epidemiology.

## Declarations

### Author contributions

ZH: conceptualization, data curation, formal analysis, validation, visualization, writing – original draft, funding acquisition. JY: methodology, validation, supervision, funding acquisition, writing – review & editing. Both authors read and approved the final manuscript.

### Data availability

The data analyzed in this study are from MIMIC-IV v3.1 and eICU-CRD v2.0. Researchers can apply for access through PhysioNet (https://physionet.org/content/mimiciv/3.1/; https://physionet.org/content/eicu-crd/2.0/) after credentialing, training and acceptance of the data use agreements.^43^ The authors cannot redistribute patient records. The code used to derive the cohort and produce the results is available in Zenodo (version 1.3.0 DOI: https://doi.org/10.5281/zenodo.24370246; concept DOI: https://doi.org/10.5281/zenodo.24214964). An earlier version of this work was posted as a preprint on medRxiv (https://doi.org/10.64898/2026.11.14.26368094).

## Acknowledgments

We thank the teams that created and maintain MIMIC-IV and eICU-CRD.

Assistance with the study: none.

Presentation: none.

## Supplemental Digital Content

Supplemental Methods (variable definitions, code lists, sensitivity analysis rules); Supplemental Figure S1 (patient flow); Supplemental Figure S2 (cross-classification heat map); Supplemental Figure S3 (Kaplan–Meier curves by unified grade [A] and agreement group [B]); Supplemental Tables S1–S6 (measurement availability and diagnosis categories, secondary outcomes and NACSELD cross-classification, first-week dynamics and post-ICU outcomes, alternative grading rules and discordant phenotypes, eICU-CRD, and subgroup analyses); STROBE and RECORD checklists.

