## Supplementary material for "Critically ill patients with cirrhosis outside the unified ACLF definition: a two-database cohort study"

Zhiqiang Hu and Jihui Ye

#### SDC, Materials and Methods

##### Cohort and time axis

MIMIC-IV v3.1 records were joined by patient, hospitalization and ICU stay. We selected the first adult ICU stay, required a cirrhosis code in its index hospitalization and required both bilirubin and INR in the grading window. Cirrhosis codes were ICD-9-CM 571.2, 571.5 and 571.6, and ICD-10-CM K70.30, K70.31, K71.7, K74.3, K74.4, K74.5, K74.60 and K74.69. Alcohol-related cirrhosis used 571.2, K70.30 or K70.31. The grading window opened 24 hours before ICU entry (inclusive) and closed at the earlier of ICU exit or 24 hours after entry (exclusive). Pre-ICU hepatic laboratories came from the index hospitalization. ICU-specific chart and treatment data contributed only when observed.

Index-hospital liver transplant used procedure codes 50.51 or 50.59 (ICD-9-CM) and 0FY00Z0, 0FY00Z1 or 0FY00Z2 (ICD-10-PCS). We used the earliest recorded transplant calendar date. Procedures on the admission date were grouped with earlier procedures as perioperative. An undated index-admission transplant would also exclude a patient, but none occurred. A transplant status code (V42.7 or Z94.4) in a prior hospitalization ended on or before entry, or in the index hospitalization without a procedure, indicated prior transplantation. A status code in the same hospitalization as a procedure did not independently indicate prior transplantation. The composite endpoint counted death or a first index-admission transplant on calendar days 1–28 after ICU entry, taking the earlier date.

MIMIC-IV death used the index-hospital death timestamp (or discharge if the death flag was positive and the timestamp absent) and linked date-only records. Timestamped deaths were measured from ICU entry and linked dates in calendar days from the entry date. We discarded dates before entry. When both sources were valid, the linked date took precedence only if it preceded the timestamp in calendar days. Deaths on or before days 28 and 90 counted. Kaplan–Meier time began at ICU entry and observations without a recorded event were administratively censored at day 90. The 24-hour landmark required confirmed survival and remaining in the ICU at 24 hours. Indeterminate date-only records were excluded in that sensitivity analysis.

eICU-CRD v2.0 selected the first recorded adult ICU stay, breaking ties with discharge year and deterministic encounter and stay identifiers. Cirrhosis codes were searched across recorded units of the hospitalization. Chronology across encounters in the same year was unavailable. The relative laboratory window and grading thresholds matched MIMIC-IV with source-specific fields and no transplant exclusion. In-hospital death was measured from recorded discharge status. Scores use both-test denominators and mortality uses known outcomes. The index-unit coding sensitivity required coding in that unit; the original complete-case sensitivity also applied original measurement rules.

#### Scores and field mappings

| Domain | MIMIC-IV source and unified rule |
|---|---|
| Hepatic | Hospital labevents. Total bilirubin item IDs 50885/53089 in mg/dL and INR item IDs 51237/51675. Each used its maximum observed value. Both ≥5 mg/dL and ≥1.5 were required. The maxima could occur at different times. |
| Renal | Serum creatinine (labevents 50912/52546, mg/dL) and ICU hourly urine output with latest measured weight. We used KDIGO changes against the lowest earlier creatinine in 48 hours or 7 days and assessed urine output over contiguous 6-, 12-, and 24-hour windows. Stage 1 indicated dysfunction, stage 2 or 3 indicated failure. New active RRT was also counted as failure. We used native treatment intervals and available maintenance-dialysis codes and excluded known maintenance RRT as a sole trigger for a new event. |
| Cerebral | ICU chart events 220739, 223900, and 223901 for eye, verbal, and motor components. The minimum total at a time when all three components were measured was used. GCS 14 indicated dysfunction and ≤13 failure. A charted unavailable verbal response due to an endotracheal tube was not assigned a numeric component. The other specifications excluded patients sedated or intubated at entry or omitted cerebral dysfunction and failure in all relevant scores. |
| Circulatory | Lowest valid arterial or noninvasive MAP from derived vital signs. MAP <65 mmHg indicated dysfunction and positive norepinephrine, epinephrine, dopamine, phenylephrine, or vasopressin administration indicated failure. Treatment indication was not ascertainable. |
| Respiratory | Derived arterial blood gases and oxygen delivery records. $PaO_2/FiO_2$ (P/F) >225 to 300 indicated dysfunction and ≤225 indicated failure. When P/F was unavailable, $SpO_2/FiO_2$ (S/F) >244 to 333 indicated dysfunction and ≤244 indicated failure. S/F used $SpO_2$ ≤97% and a $FiO_2$ record no more than one hour earlier. Recorded invasive ventilation indicated failure under the operational rule. The clinical indication was unavailable. |
| Peak lactate | Hospital labevents items 50813, 52442, and 53154. Highest positive in-window measurement in mmol/L. We report the number measured and summarize available values. |
| Other laboratory inputs | Platelets (51265/53189), serum sodium (50983/52623), and albumin (50862/53085) came from labevents. We used the minimum in-window value for each. |

KDIGO references used only earlier creatinine, extracted from seven days before entry through the earlier of ICU exit or the extraction cap (48 hours after entry in MIMIC-IV and 24 hours in eICU-CRD). The 48-hour and seven-day windows used prior minima. Earlier MIMIC-IV encounters could provide reference values. Grade 1 required both hepatic thresholds, extrahepatic dysfunction and no extrahepatic failure. Grades 2 and 3 required both thresholds and one or at least two extrahepatic failures, respectively. Other patients had grade 0. Clinical A/B type and decompensation timing could not be assigned.

EASL-CLIF failure flags were bilirubin ≥12 mg/dL, creatinine ≥2 mg/dL or RRT, GCS ≤13, INR ≥2.5, vasopressor support, and P/F ≤200 or S/F ≤214 when P/F was missing, or invasive ventilation. CLIF-C OF uses three bands per domain, with a renal score of 2 for creatinine 2 to <3.5 mg/dL unless RRT applies. EASL-CLIF grades 2 and 3 required two and at least three failure flags. Grade 1 required isolated renal failure or another failure with qualifying renal or cerebral dysfunction; for isolated cerebral failure, creatinine had to be 1.5 to <2 mg/dL. GCS 14 represented cerebral dysfunction. NACSELD positivity required ≥2 of RRT, GCS ≤13, invasive ventilation or documented BiPAP, and vasopressor use. Recorded vasopressors proxied circulatory failure without adjudicating indication or persistent hypotension.

MELD 3.0 and MELD-Na used the published input bounds, sodium and dialysis rules. Missing creatinine, sodium and albumin were assigned 1 mg/dL, 137 mmol/L and 3.5 g/dL, respectively. Missing sex did not add the female coefficient; dialysis used observable treatment in the seven days before window end. Historical MELD-Na followed OPTN policy 9.1.D. Initial MELD was $3.78 \times \ln(\text{bilirubin}) + 11.20 \times \ln(\text{INR}) + 9.57 \times \ln(\text{creatinine}) + 6.43$, with bilirubin and creatinine in mg/dL. Inputs were floored at 1; creatinine was capped at 4 mg/dL and set to 4 when the dialysis rule applied. The initial score was rounded and bounded to 6–40. Above 11, MELD-Na added $(1.32 - 0.033 \times \text{initial MELD}) \times (137 - \text{sodium})$, bounding sodium at 125–137 mmol/L and rounding and bounding the score to 6–40.

Classic SOFA used six domains, requiring respiratory support at the P/F assessment for scores 3–4. The renal component incorporated creatinine and available urine output over approximately 24 hours. Unmeasured nonhepatic domains used the normal band. In eICU-CRD, we used contemporaneous GCS components, arterial oxygenation paired to the last eligible $FiO_2$ within four hours (S/F if P/F unavailable), and recorded respiratory support, MAP, vasoactive drugs and dialysis. Dose-based SOFA and the MELD dialysis override were limited by missing dose or treatment duration. eICU-CRD bootstrap intervals used 2,000 patient-level resamples. Each score's 28-day Harrell C treated deaths at the day-28 cutoff as comparable with patients without a death by that cutoff, omitted ties in death time and assigned half credit for score ties.

#### Supplemental analyses

The eICU-CRD and MIMIC-IV first-week cohorts included EASL-CLIF-positive patients with unified grade 0 at baseline, including those with both hepatic thresholds but no qualifying extrahepatic finding. Days were half-open 24-hour blocks. Assessment ran from 24 hours (start of D2) to before hour 168 (end of D7). Both qualifying measurements were required within the same day and before a competing event. The later of the first qualifying bilirubin (≥5 mg/dL) and INR (≥1.5) draws marked the event in that block; the first such time was used.

Follow-up began at ICU entry. Live ICU exit, ICU death and timed transplant competed in MIMIC-IV. Transplants were timed at the start of their recorded calendar date. Death took precedence if tied with transplant. Live ICU exit and ICU death competed in eICU-CRD. Vital status unknown at ICU exit was right-censored at exit, and transplant timing was unavailable. Aalen–Johansen used a common risk set for ties. First threshold events strictly before day-end and exits through day-end contributed to daily counts, including exits before hour 24. Pointwise 95% CIs used Wilson intervals on the starting denominator in MIMIC-IV and 2,000 patient-level percentile bootstrap resamples in eICU-CRD.

Among threshold-positive patients, extrahepatic domains were reassessed on the same ICU-relative day and within ±24 hours of the first threshold time. Windows ended at ICU exit, death, transplant or hour 168; the same-day analysis also ended at the next day boundary. We distinguished qualifying findings, assessed domains without a finding and no effective domain evidence. The MIMIC-IV day-7 landmark assessed deaths through day 28 from ICU entry in patients alive at hour 168 with a documented threshold or at least one evaluable day without it. Patients already discharged from the ICU could qualify. No paired days or indeterminate survival at 168 hours excluded a patient from the comparison. This was a descriptive, unadjusted analysis.

Baseline sensitivity analyses changed cerebral thresholds or removed the cerebral domain, restricted respiratory failure to oxygenation, restricted to patients not intubated at entry, changed transplant exclusions or analyzed the composite, varied ICU windows, or excluded elective admissions (admission type ELECTIVE). Joint complete cases required creatinine, albumin, sodium, platelets, GCS, arterial P/F, MAP, usable KDIGO staging or new RRT, and recorded sex. Three additional post hoc variants kept the main cohort, EASL-CLIF grades and 28-day outcomes fixed: remove GCS 14 as dysfunction but retain ≤13 as failure; require bilirubin ≥5 and INR ≥1.5 within 12 hours of each other, inclusive; and assign grade 0 to patients with ≥2 extrahepatic dysfunctions and no failure who would otherwise have grade 1. Patients without a qualifying pair remained in the cohort.

The alcohol-related cirrhosis and entry-intubation subgroups used the original definitions. Entry intubation was invasive ventilation overlapping the first hour or a contemporaneous endotracheal tube indicator. Entry sedation was a sedative infusion spanning entry. For the updated infection subgroup, we classified a patient as having suspected infection around ICU admission if the earlier timestamp of a qualifying culture–antibiotic pair fell from 24 hours before ICU entry inclusive to 24 hours after entry exclusive. The second element could fall outside that window.

The MIMIC-IV derived pairing logic linked microbiology to the same patient and used antibiotic starts from the index hospitalization, including pre-ICU records without an ICU stay identifier. With exact times, a culture preceding an antibiotic start qualified if the start was within 72 hours. If the antibiotic came first, the culture qualified within 24 hours. For date-only cultures, it used calendar days, allowing up to three days from culture to antibiotic or one day from antibiotic to culture. Eligible culture dates without a clock time were set to midnight for the anchor. The proxy includes qualifying pairs involving prophylactic antibiotics, and recorded antibiotic starts can come from prescriptions. A qualifying pair indicates suspected infection. We classified infections and decompensating events from index-hospitalization diagnoses at any coding position. The infection and sepsis codes used the infection category in the principal-diagnosis map below, excluding ICD-10-CM J69. Diagnosis codes could not establish onset.

For patients whose first event was live ICU exit before hour 168, we described death by day 28 by recorded location and hospice disposition using the index ICU and hospital discharge records. Date-only records at discharge boundaries could leave death location unresolved.

Fresh frozen plasma (FFP) was identified from qualifying eMAR records for FFP or fresh frozen plasma with an administered, started, new bag, given or partial administered status, and from ICU inputevents (items 220970, 226367 or 227072 with positive amount, excluding rewritten or canceled entries). We counted each patient once if eMAR chart time or infusion start time fell from 24 hours before ICU entry inclusive to 24 hours after entry exclusive. A recorded administration alone does not establish if it preceded INR sampling.

Coded decompensation matched normalized whole ICD-CM codes in any diagnosis position in the index hospitalization. These codes could not establish acuity or onset relative to ICU entry. The first-listed index-hospital diagnosis (seq_num = 1) approximated the principal admission condition. Decimal-free, upper-case codes were assigned by the first matching category in the code map, with explicit gastrointestinal bleeding and hepatic encephalopathy taking precedence. Infection included pneumonia; the remaining unmatched codes were Other. We excluded cardiovascular, postoperative and neoplasm categories in the relevant sensitivity analyses.

##### Coded decompensation definitions

| Category | ICD-9-CM | ICD-10-CM |
|---|---|---|
| Ascites | 789.50, 789.59 | R18.8, K70.31, K70.11, K71.51 |
| Hepatic encephalopathy and historical hepatic coma codes | 572.2 | K76.82, K70.41, K71.11, K72.01, K72.11, K72.91 |
| Bleeding esophageal varices | 456.0, 456.20 | I85.01, I85.11 |
| Hepatorenal syndrome | 572.4 | K76.7 |
| Spontaneous bacterial peritonitis | 567.23 | K65.2 |
| Jaundice | 782.4 | R17 |

##### Principal diagnosis code map

| Principal diagnosis category | ICD-9-CM alternatives | ICD-10-CM alternatives |
|---|---|---|
| Gastrointestinal bleeding | 4560, 45620, 578[019], 5307, 531[0246]\d?, 532[0246]\d?, 533[0246]\d?, 534[0246]\d?, 535[0-6]1 | I8501, I8511, I864, K920, K921, K922, K226, K2[5-8][0246], K29\d*1, K57\d*[13] |
| Hepatic encephalopathy | 5722 | K7682 |
| Liver-related | 570, 571[0-9]\d*, 572[0348], 573[0-9]\d*, 7895[09]?, 56723, 4561, 45621 | K70\w*, K71\w*, K72\w*, K73\w*, K74\w*, K75\w*, K76\w*, R18\w*, K652, I859[01], I8500, I8510 |
| Infection or sepsis | 00[1-9]\d*, 01[0-8]\d*, 03[0-8]\d*, 04[0-1]\d*, 08[0-8]\d*, 480\d*, 48[1-8]\d*, 590\d*, 5990, 567\d*, 9959[12], 78552 | A0[0-9]\w*, A1[5-9]\w*, A3[0-9]\w*, A4[0-9]\w*, B3[4-5]\w*, B37\w*, J1[0-8]\w*, J69\w*, N10, N12, N390, K65\w*, R652[01], U071 |
| Respiratory | 49[0-6]\d*, 51[0-9]\d*, 7860\d*, 7990[12]? | J4[0-7]\w*, J8[0-6]\w*, J9[0-9]\w*, R06\w*, R0902 |
| Renal | 58[4-8]\d*, 5724 | N1[7-9]\w*, N99[01]\w*, K767 |
| Postoperative | 996[6-7]\d*, 997\d*, 998\d*, V54\d*, V58[347]\d* | T8[0-5]\w*, Z4[78]\w*, K91\w*, J95\w*, I97\w* |
| Cardiovascular | 39\d{1,3}, 4[0-5]\d{1,3}, 785[0149]\d* | I[0-9]\w*, R00\w*, R07[29]\w* |
| Neoplasm | 1[4-9]\d{1,3}, 2[0-3]\d{1,3} | C[0-9]\w*, D[0-4]\w* |
| Other | All remaining codes | All remaining codes |

Supplemental reference. OPTN and UNOS. Enhancing Liver Distribution. Policy Notice, 2017, section 9.1.D. https://www.hrsa.gov/sites/default/files/hrsa/optn/liver_policynotice_20171221.pdf

#### SDC, Figure S1

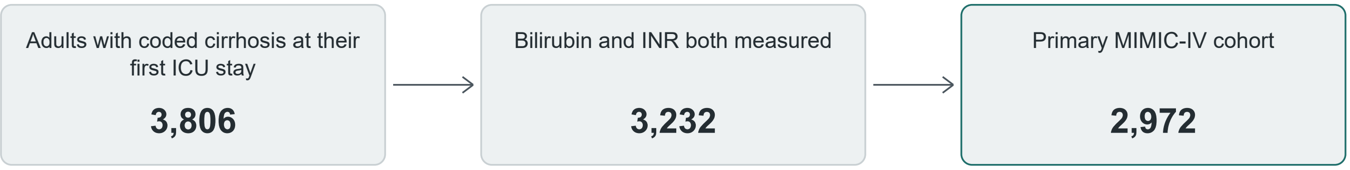


**Selection of the primary MIMIC-IV cohort.** Of 3,806 eligible patients, 574 lacked one or both baseline laboratory measurements. An additional 260 were excluded for prior transplantation or index-admission transplantation on or before ICU admission. No index-admission transplant had unknown timing. See Supplemental Table S5 for replication flow and hospital counts.

#### SDC, Figure S2

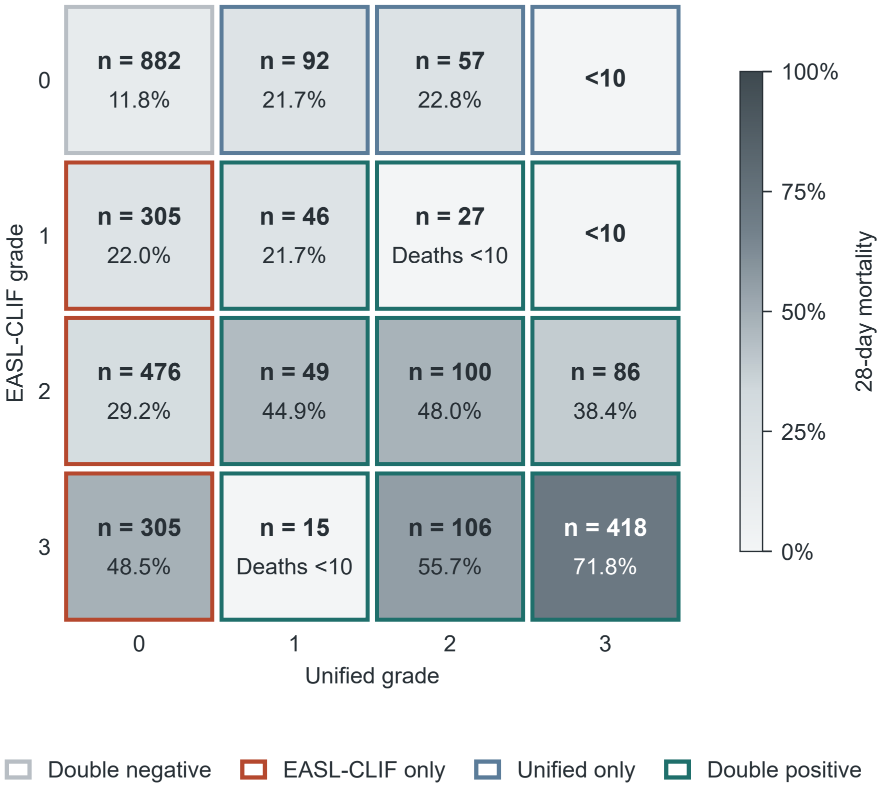


**Cross-classification and 28-day mortality.** Rows show EASL-CLIF grades and columns show unified grades in the primary MIMIC-IV cohort. Counts indicate the number of patients. Shading and percentages represent 28-day mortality. Borders identify the four agreement groups. Cells with fewer than 10 patients or deaths omit mortality percentages and use a neutral fill.

### SDC, Figure S3

**A**

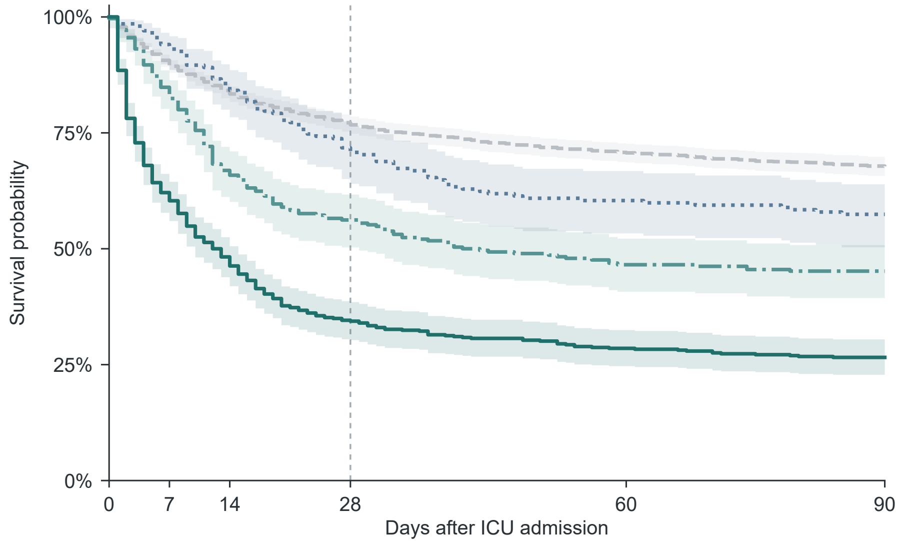


**Number at risk**

| | 0 | 7 | 14 | 28 | 60 | 90 |
|---|---|---|---|---|---|---|
| Grade 0 | 1,968 | 1,762 | 1,645 | 1,515 | 1,393 | 1,335 |
| Grade 1 | 202 | 188 | 173 | 145 | 122 | 116 |
| Grade 2 | 290 | 239 | 191 | 163 | 135 | 131 |
| Grade 3 | 512 | 310 | 238 | 177 | 146 | 136 |

**B**

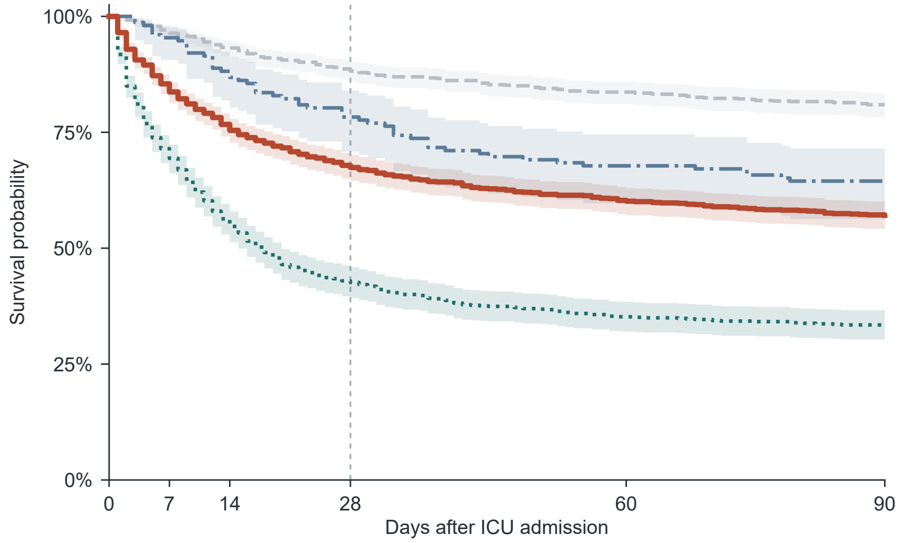


**Number at risk**

| | 0 | 7 | 14 | 28 | 60 | 90 |
|---|---|---|---|---|---|---|
| Double negative | 882 | 850 | 823 | 781 | 738 | 714 |
| **EASL-CLIF only** | 1,086 | 912 | 822 | 734 | 655 | 621 |
| Unified only | 152 | 145 | 133 | 119 | 103 | 98 |
| Double positive | 852 | 592 | 469 | 366 | 300 | 285 |

**Kaplan–Meier survival by unified grade and agreement group.** (A) Unified grades 0–3. (B) Agreement groups defined by EASL-CLIF and unified grades. Both panels use the primary MIMIC-IV cohort and show all-cause survival through day 90. Shading gives pointwise log–log Greenwood 95% confidence intervals. Counts are at risk immediately before the indicated day. The dashed line marks day 28.

### Supplemental tables

Counts below 10 are displayed as <10. Their percentages and intervals are omitted. A dash indicates a result unavailable or reported separately.

#### SDC, Table S1. Measurement availability in MIMIC-IV

| Variable | Measured | Missing |
|---|---|---|
| Bilirubin | 2,972 (100.0%) | <10 |
| INR | 2,972 (100.0%) | <10 |
| Glasgow Coma Scale | 2,405 (80.9%) | 567 (19.1%) |
| Arterial P/F | 987 (33.2%) | 1,985 (66.8%) |
| Albumin | 2,257 (75.9%) | 715 (24.1%) |
| Sodium | 2,972 (100.0%) | <10 |
| Creatinine | 2,972 (100.0%) | <10 |
| Platelet count | 2,969 (99.9%) | <10 |
| Mean arterial pressure | 2,963 (99.7%) | <10 |
| Lactate | 2,281 (76.7%) | 691 (23.3%) |

The Supplemental Methods give the corresponding variable and diagnosis definitions. Tabulations use the primary cohort denominator of 2,972.

#### SDC, Table S2. Baseline prevalence, secondary outcomes and comparison scores

##### A. Baseline prevalence

| Definition | Positive, N | Proportion (95% CI) |
|---|---|---|
| Unified | 1,004/2,972 | 33.8% (32.1%–35.5%) |
| EASL-CLIF | 1,938/2,972 | 65.2% (63.5%–66.9%) |
| NACSELD | 889/2,972 | 29.9% (28.3%–31.6%) |

##### B. Secondary mortality

| Unified grade | 90-day mortality | In-hospital mortality |
|---|---|---|
| 0 | 634/1,968 (32.2%; 30.2%–34.3%) | 368/1,968 (18.7%; 17.0%–20.5%) |
| 1 | 86/202 (42.6%; 36.0%–49.5%) | 44/202 (21.8%; 16.6%–28.0%) |
| 2 | 159/290 (54.8%; 49.1%–60.5%) | 118/290 (40.7%; 35.2%–46.4%) |
| 3 | 376/512 (73.4%; 69.4%–77.1%) | 319/512 (62.3%; 58.0%–66.4%) |

##### C. Share of deaths classified negative

| Definition | MIMIC-IV day 28 | MIMIC-IV day 90 |
|---|---|---|
| Unified | 458/980 (46.7%; 43.6%–49.9%) | 634/1,255 (50.5%; 47.8%–53.3%) |
| EASL-CLIF | 137/980 (14.0%; 11.9%–16.3%) | 222/1,255 (17.7%; 15.7%–19.9%) |
| NACSELD | 539/980 (55.0%; 51.9%–58.1%) | 743/1,255 (59.2%; 56.5%–61.9%) |

##### D. Supplementary discrimination measures

| Score | AUC | Harrell C |
|---|---|---|
| NACSELD positive | 0.613 (0.594 to 0.630) | 0.603 (0.587 to 0.617) |
| MELD 3.0 | 0.758 (0.740 to 0.776) | 0.724 (0.709 to 0.739) |
| MELD-Na | 0.757 (0.738 to 0.775) | 0.724 (0.708 to 0.739) |
| SOFA | 0.760 (0.742 to 0.778) | 0.733 (0.717 to 0.747) |

Mortality and prevalence intervals use Wilson proportions. Discrimination intervals come from 2,000 paired bootstrap resamples. AUCs for the matched-resolution comparisons and CLIF-C OF reference are in Table 3.

##### E. Unified grade × NACSELD cross-classification

| Unified ACLF | NACSELD ACLF | MIMIC-IV day-28 deaths | eICU-CRD in-hospital deaths |
|---|---|---|---|
| Negative | Negative | 269/1,464 (18.4%, 16.5%–20.4%) | 87/609 (14.3%, 11.7%–17.3%) |
| Negative | Positive | 189/504 (37.5%, 33.4%–41.8%) | 44/129 (34.1%, 26.5%–42.6%) |
| Positive | Negative | 270/619 (43.6%, 39.8%–47.6%) | 74/182 (40.7%, 33.8%–47.9%) |
| Positive | Positive | 252/385 (65.5%, 60.6%–70.0%) | 48/82 (58.5%, 47.7%–68.6%) |

Unified grades 1–3 are positive. Cells show deaths/known outcomes (% and Wilson 95% CI). The four eICU-CRD classification counts in table order were 613, 130, 183 and 82 before restriction to known hospital discharge status. All 2,972 MIMIC-IV patients had 28-day outcome data.

#### SDC, Table S3. First-week course in patients positive by EASL-CLIF alone

##### A. First events and follow-up status

| First event or observation status at 168 hours | MIMIC-IV | eICU-CRD |
|---|---|---|
| Starting EASL-CLIF-only cohorts | 1,086 | 343 |
| First observed liver-failure threshold (95% CI) | 134/1,086 (12.3%; 10.5%–14.4%) | 33/343 (9.6%; 6.7%–12.5%) |
| Discharged alive before threshold observed | 645 | 210 |
| Died in ICU before threshold observed | 118 | 41 |
| Transplant before threshold observed | <10 | — |
| ICU exit of unknown vital status, censored at exit | — | <10 |
| No first event by 168 hours | 186 | 59 |

The row for a first observed threshold gives events/starting cohort (Aalen–Johansen estimate and pointwise 95% CI through hour 168). Intervals are Wilson in MIMIC-IV and patient-level percentile bootstrap in eICU-CRD. A dash indicates an event type unavailable or inapplicable.

##### B. Daily states and observation after the baseline grading window

| Database and day | Threshold observed | Live ICU exit | ICU death or other exit | Paired below threshold | No paired tests | Cumulative incidence |
|---|---|---|---|---|---|---|
| MIMIC-IV D2 | 68 | 266 | 76 | 492 | 184 | 6.3% |
| MIMIC-IV D3 | 103 | 403 | 91 | 307 | 182 | 9.5% |
| MIMIC-IV D4 | 117 | 501 | 97 | 229 | 142 | 10.8% |
| MIMIC-IV D5 | 120 | 571 | 106 | 171 | 118 | 11.0% |
| MIMIC-IV D6 | 130 | 610 | 111 | 140 | 95 | 12.0% |
| MIMIC-IV D7 | 134 | 645 | 121 | 105 | 81 | 12.3% |
| eICU-CRD D2 | 16 | 82 | 28 | 80 | 137 | 4.7% |
| eICU-CRD D3 | 23 | 134 | 33 | 52 | 101 | 6.7% |
| eICU-CRD D4 | 26 | 159 | 35 | 44 | 79 | 7.6% |
| eICU-CRD D5 | 27 | 182 | 39 | 32 | 63 | 7.9% |
| eICU-CRD D6 | 31 | 198 | 41 | 16 | 57 | 9.0% |
| eICU-CRD D7 | 33 | 210 | 41 | 22 | 37 | 9.6% |

Paired-test categories describe patients still under observation at each day-end. The pooled other-exit category includes ICU death and transplantation in MIMIC-IV or ICU death and ICU exit of unknown vital status in eICU-CRD. Unknown eICU-CRD exits remain in the displayed counts and are censored in the Aalen–Johansen estimates.

##### C. Extrahepatic domains among those with an observed threshold

| Database and window | Threshold observed | Qualifying extrahepatic finding | Examined without qualifying finding | No effective domain evidence |
|---|---|---|---|---|
| MIMIC-IV, same day | 134 | 126 | <10 | <10 |
| MIMIC-IV, ±24 hours | 134 | 128 | <10 | <10 |
| eICU-CRD, same day | 33 | 30 | <10 | <10 |
| eICU-CRD, ±24 hours | 33 | 30 | <10 | <10 |

##### D. Descriptive MIMIC-IV day-7 landmark

| MIMIC-IV day-7 status | Patients | 28-day deaths (95% CI) |
|---|---|---|
| Alive at day 7, threshold observed | 106 | 41/106 (38.7%; 30.0%–48.2%) |
| Alive at day 7, paired tests but threshold not observed | 513 | 98/513 (19.1%; 15.9%–22.7%) |
| Alive at day 7, no paired tests | 290 | — |
| Survival at day 7 not established | 177 | — |

Day-28 mortality is from ICU entry, among the two evaluable day-7 survivor groups.

##### E. Death location and disposition among 645 patients with live ICU exit as first event

| Outcome or disposition | All EASL-CLIF only, N = 1,086 | Live exit first, N = 645 |
|---|---|---|
| Death by day 28, n (% and Wilson 95% CI) | 354 (32.6%, 29.9%–35.4%) | 112 (17.4%, 14.6%–20.5%) |
| Death during index ICU stay | 197 | — |
| Death after index ICU exit during index hospitalization | 78 | 51 |
| Death after index-hospital discharge | 71 | 55 |
| Other or unresolved timing/location | <10 | <10 |
| Discharged alive to hospice, n (% and Wilson 95% CI) | — | 41 (6.4%, 4.7%–8.5%) |
| Discharged alive to hospice within 28 days, n | — | 35 |

Death locations count deaths through day 28 after ICU entry. For the live-exit group, deaths outside the two classifiable post-exit categories are combined. Hospice is a discharge disposition and can overlap with 28-day death.

#### SDC, Table S4. Sensitivity to alternative classification and cohort rules

##### A. Agreement-group counts and deaths across grading rules

| Analysis | N | Both− | EASL-CLIF only | Unified only | Both+ | EASL-CLIF only: 28-day death, n/N (%, 95% CI) |
|---|---|---|---|---|---|---|
| Primary analysis | 2,972 | 882 | 1,086 | 152 | 852 | 354/1,086 (32.6%; 29.9%–35.4%) |
| Excluded sedation or intubation at ICU entry | 2,304 | 766 | 761 | 132 | 645 | 242/761 (31.8%; 28.6%–35.2%) |
| Cerebral domain omitted | 2,972 | 1,075 | 951 | 197 | 749 | 333/951 (35.0%; 32.0%–38.1%) |
| Joint complete cases | 447 | 45 | 210 | <10 | 187 | 63/210 (30.0%; 24.2%–36.5%) |
| First 48 hours after ICU entry | 2,971 | 804 | 1,098 | 135 | 934 | 334/1,098 (30.4%; 27.8%–33.2%) |
| First 24 hours after ICU entry | 2,802 | 855 | 1,005 | 146 | 796 | 332/1,005 (33.0%; 30.2%–36.0%) |
| Alive and in ICU at 24 hours | 2,449 | 627 | 969 | 116 | 737 | 304/969 (31.4%; 28.5%–34.4%) |
| No transplant exclusion | 3,232 | 928 | 1,181 | 163 | 960 | 356/1,181 (30.1%; 27.6%–32.8%) |
| Elective admissions excluded | 2,943 | 874 | 1,067 | 152 | 850 | 352/1,067 (33.0%; 30.2%–35.9%) |
| Oxygenation-only respiratory failure | 2,972 | 1,016 | 957 | 175 | 824 | 334/957 (34.9%; 31.9%–38.0%) |
| Not intubated at ICU entry | 2,308 | 767 | 763 | 132 | 646 | 242/763 (31.7%; 28.5%–35.1%) |
| Any index-admission transplant or prior transplant status excluded | 2,905 | 878 | 1,074 | 152 | 801 | 354/1,074 (33.0%; 30.2%–35.8%) |
| Coded decompensation | 1,883 | 423 | 616 | 114 | 730 | 211/616 (34.3%; 30.6%–38.1%) |
| Coded decompensation with diagnosis exclusions | 1,665 | 345 | 525 | 104 | 691 | 175/525 (33.3%; 29.4%–37.5%) |
| Principal diagnosis exclusions only | 2,438 | 663 | 854 | 129 | 792 | 272/854 (31.9%; 28.8%–35.1%) |
| GCS 14 omitted as dysfunction | 2,972 | 894 | 1,101 | 140 | 837 | 357/1,101 (32.4%; 29.7%–35.2%) |
| Hepatic thresholds within 12 hours | 2,972 | 884 | 1,089 | 150 | 849 | 355/1,089 (32.6%; 29.9%–35.4%) |

| Analysis | N | Both− | EASL-CLIF only | Unified only | Both+ | EASL-CLIF only: 28-day death, n/N (%, 95% CI) |
|---|---|---|---|---|---|---|
| Grade 0 for multiple dysfunctions without failure | 2,972 | 912 | 1,147 | 122 | 791 | 381/1,147 (33.2%; 30.6%–36.0%) |

All rows use all-cause death within 28 days of ICU entry. The principal diagnosis exclusions cover cardiovascular, postoperative and neoplasm categories. The separate cerebral checks either removed GCS 14 as dysfunction or omitted the whole cerebral domain.

##### B. Transplant analysis

| Analysis | N | Both− | EASL-CLIF only | Unified only | Both+ | EASL-CLIF only: death/transplant, n/N (%, 95% CI) |
|---|---|---|---|---|---|---|
| Death or transplant by day 28 | 2,972 | 882 | 1,086 | 152 | 852 | 365/1,086 (33.6%; 30.9%–36.5%) |

Of 306 patients with a transplant in the index hospitalization before analytic exclusions, 239 had the procedure on or before ICU entry and 67 after ICU entry. The composite counted death or an incident transplant on calendar days 1–28. Any index-admission transplant or prior transplant status excluded and No transplant exclusion are separately shown in panel A.

##### C. Distribution of deaths in selected restrictions (S4 continued)

| Respiratory or admission restriction | Deaths classified unified negative, day 28 | Deaths classified EASL-CLIF negative, day 28 | Unified negative, day 90 | EASL-CLIF negative, day 90 |
|---|---|---|---|---|
| Elective admissions excluded | 456/977 (46.7%; 43.6%–49.8%) | 137/977 (14.0%; 12.0%–16.3%) | 631/1,250 (50.5%; 47.7%–53.2%) | 222/1,250 (17.8%; 15.7%–20.0%) |
| Oxygenation-only failure | 461/980 (47.0%; 43.9%–50.2%) | 167/980 (17.0%; 14.8%–19.5%) | 637/1,255 (50.8%; 48.0%–53.5%) | 263/1,255 (21.0%; 18.8%–23.3%) |
| Not intubated at ICU entry | 324/703 (46.1%; 42.4%–49.8%) | 108/703 (15.4%; 12.9%–18.2%) | 464/928 (50.0%; 46.8%–53.2%) | 181/928 (19.5%; 17.1%–22.2%) |

Denominators are all deaths in the corresponding cohort and time horizon. Code-based decompensation could not establish onset before ICU admission.

##### D. Mortality and grade changes in post hoc baseline variants

| Baseline grading rule | Both negative | EASL-CLIF only | Unified only | Both positive | Unified grade changed |
|---|---|---|---|---|---|
| Primary analysis | 104/882 (11.8%; 9.8%–14.1%) | 354/1,086 (32.6%; 29.9%–35.4%) | 33/152 (21.7%; 15.9%–28.9%) | 489/852 (57.4%; 54.0%–60.7%) | — |
| GCS 14 omitted as dysfunction | 105/894 (11.7%; 9.8%–14.0%) | 357/1,101 (32.4%; 29.7%–35.2%) | 32/140 (22.9%; 16.7%–30.5%) | 486/837 (58.1%; 54.7%–61.4%) | 27 |
| Hepatic thresholds within 12 hours | 105/884 (11.9%; 9.9%–14.2%) | 355/1,089 (32.6%; 29.9%–35.4%) | 32/150 (21.3%; 15.5%–28.6%) | 488/849 (57.5%; 54.1%–60.8%) | <10 |
| Multiple dysfunctions without failure assigned grade 0 | 113/912 (12.4%; 10.4%–14.7%) | 381/1,147 (33.2%; 30.6%–36.0%) | 24/122 (19.7%; 13.6%–27.6%) | 462/791 (58.4%; 54.9%–61.8%) | 91 |

The four agreement-group cells report 28-day all-cause deaths over known outcomes and Wilson 95% intervals. The rightmost column compares the corresponding unified grades with those from the primary analysis.

At least one bilirubin–INR pair no more than 12 hours apart was available for 2,962 of 2,972 patients. A pair meeting both hepatic thresholds was available for 1,051 patients.

##### E. Organ findings in patients positive by only one classification

| EASL-CLIF positive only, meets both hepatic thresholds | n | Unified positive only | n |
|---|---|---|---|
| All: no qualifying extrahepatic finding under unified grading | 19 | All: met both hepatic thresholds | 152 |
| EASL-CLIF grades 1 / 2 / 3 | <10 / 14 / <10 | Unified grades 1 / 2 / 3 | 92 / 57 / <10 |
| Grade 1: isolated renal failure | <10 | No EASL-CLIF organ failure | 78 |
| Grade 1: liver or coagulation failure with creatinine 1.5 to <2 mg/dL | <10 | One EASL-CLIF nonrenal failure without other grade-1 criteria | 74 |
| Grade 2: liver and coagulation failure | 13 | Isolated circulatory dysfunction | 45 |
| Grade 2: renal and coagulation failure | <10 | Isolated cerebral dysfunction | 12 |
| Grade 3: renal, liver and coagulation failure | <10 | Renal and circulatory dysfunction | 12 |
| | | Circulatory dysfunction and respiratory failure | 12 |

The 19 patients had both hepatic laboratory thresholds but no qualifying recorded extrahepatic finding. INR contributes to the unified hepatic criterion rather than a separate coagulation domain. Among the 152 unified-only patients, the four organ patterns listed each occurred in at least 10 patients. Grade, comparator-failure and pattern rows summarize the same patients in different ways.

#### SDC, Table S5. Replication in eICU-CRD

##### A. Cohort and hospital coverage

| Step | Patients | Hospitals |
|---|---|---|
| First recorded adult ICU stay | 132,654 | 208 |
| Adults with coded cirrhosis | 1,247 | 151 |
| Bilirubin measured | 1,133 | 147 |
| Bilirubin and INR measured | 1,008 | 139 |
| Both tests and known outcome | 1,002 | 138 |

##### B. Definition positivity and share of deaths classified negative

| Definition | All scored | Complete cases | All scored: share of recorded deaths among negatives |
|---|---|---|---|
| Unified | 265/1,008 (26.3%; 23.7%–29.1%) | 50/154 (32.5%; 25.6%–40.2%) | 131/253 (51.8%; 45.6%–57.9%) |
| EASL-CLIF | 550/1,008 (54.6%; 51.5%–57.6%) | 130/154 (84.4%; 77.9%–89.3%) | 44/253 (17.4%; 13.2%–22.5%) |
| NACSELD | 212/1,008 (21.0%; 18.6%–23.7%) | 94/154 (61.0%; 53.2%–68.4%) | 161/253 (63.6%; 57.5%–69.3%) |

##### C. Grade- and agreement-specific in-hospital mortality (S5 continued)

| Grade or agreement group | Scored | In-hospital mortality |
|---|---|---|
| Unified grade 0 | 743 | 131/738 (17.8%; 15.2%–20.7%) |
| Unified grade 1 | 80 | 18/79 (22.8%; 14.9%–33.2%) |
| Unified grade 2 | 82 | 42/82 (51.2%; 40.6%–61.7%) |
| Unified grade 3 | 103 | 62/103 (60.2%; 50.5%–69.1%) |
| Both negative | 400 | 32/396 (8.1%; 5.8%–11.2%) |
| EASL-CLIF only | 343 | 99/342 (28.9%; 24.4%–34.0%) |
| Unified only | 58 | 12/57 (21.1%; 12.5%–33.3%) |
| Both positive | 207 | 110/207 (53.1%; 46.3%–59.8%) |

##### D. Index-unit coding and complete-case sensitivity

| Analysis | N | Both− | EASL-CLIF only | Unified only | Both+ | EASL-CLIF only: in-hospital death, n/N (%, 95% CI) |
|---|---|---|---|---|---|---|
| Index ICU unit coding only | 984 | 389 | 336 | 58 | 201 | 99/335 (29.6%; 24.9%–34.6%) |
| Index ICU unit coding, complete cases | 154 | 17 | 87 | <10 | 43 | 29/86 (33.7%; 24.6%–44.2%) |

The all-scored cohort consisted of 1,008 patients from 139 hospitals. Mortality analyses used the 1,002 patients (138 hospitals) with a recorded discharge status. Wilson intervals are based on known outcomes. Unified grade 0 included 343/550 (62.4%) EASL-CLIF-positive patients in the all-scored cohort.

#### SDC, Table S6. Subgroup comparisons

| Subgroup | Cohort N | Grade 0 day-28 deaths | Grade 3 day-28 deaths | Unified grade AUC | EASL-CLIF grade AUC |
|---|---|---|---|---|---|
| No alcohol-related code | 1,294 | 253/1,002 (25.2%; 22.7%–28.0%) | 105/149 (70.5%; 62.7%–77.2%) | 0.627 | 0.718 |
| Alcohol-related code | 1,678 | 205/966 (21.2%; 18.8%–23.9%) | 231/363 (63.6%; 58.6%–68.4%) | 0.693 | 0.748 |
| No qualifying culture–antimicrobial pair | 763 | 98/552 (17.8%; 14.8%–21.2%) | 56/78 (71.8%; 61.0%–80.6%) | 0.684 | 0.746 |
| Suspected infection around ICU admission | 2,209 | 360/1,416 (25.4%; 23.2%–27.8%) | 280/434 (64.5%; 59.9%–68.9%) | 0.657 | 0.724 |
| Not intubated at entry | 2,308 | 324/1,530 (21.2%; 19.2%–23.3%) | 217/330 (65.8%; 60.5%–70.7%) | 0.668 | 0.740 |
| Intubated at entry | 664 | 134/438 (30.6%; 26.5%–35.1%) | 119/182 (65.4%; 58.2%–71.9%) | 0.655 | 0.706 |

Alcohol-related cirrhosis, infection and intubation at entry were prespecified subgroup factors. The endpoint was death within 28 days of ICU entry.

The infection strata were recalculated post hoc. The updated definition used the earlier timestamp of a qualifying culture–antimicrobial pair from 24 hours before to 24 hours after ICU admission (right endpoint excluded). Version 1 of the preprint used the later timestamp in a narrower window starting 48 hours before ICU admission and ending just before hour +1. AUCs are for the two ordinal grades, and mortality intervals are Wilson 95% confidence intervals.

#### STROBE cohort reporting checklist

Critically ill patients with cirrhosis outside the unified ACLF definition: a two-database cohort study

Manuscript page numbers refer to the medRxiv manuscript PDF. SDC pages refer to this supplement.

| Item | Reporting item | Location and reporting note |
|---|---|---|
| 1(a) | Study design in the title or abstract | Manuscript p. 1 specifies a two-database cohort study. Manuscript p. 2 and manuscript pp. 3–4 identify the data sources. |
| 1(b) | Balanced abstract | Manuscript p. 2 |
| 2 | Background and rationale | Manuscript p. 3 |
| 3 | Objectives and planned hypotheses, where applicable | Manuscript p. 3 |
| 4 | Design features early in the paper | Manuscript pp. 3–4 |
| 5 | Setting, periods and observation dates | Manuscript pp. 3–4 |
| 6(a) | Eligibility, selection and follow-up | Manuscript pp. 3–5; SDC pp. 1–4 (Supplemental Methods); SDC p. 5 (Figure S1). |
| 6(b) | Matching and group sizes if matched | Not applicable. No matching was used. |
| 7 | Variable definitions and criteria | Manuscript pp. 4–5; SDC pp. 1–4 (Supplemental Methods). |
| 8 | Data sources and comparable measurement | Manuscript pp. 3–5; SDC pp. 1–4 (Supplemental Methods). |
| 9 | Measures to address bias | Manuscript pp. 4–5; Manuscript pp. 7–8; Manuscript pp. 9–10. |
| 10 | How the study size was obtained | Manuscript p. 6; SDC p. 5 (Figure S1). All eligible cases in the stated data sources were used. |
| 11 | Handling quantitative variables and thresholds | Manuscript pp. 4–5; SDC pp. 1–4 (Supplemental Methods). |
| 12(a) | Statistical methods and handling of confounding | Manuscript p. 5. The prognostic and reclassification comparisons are descriptive. |
| 12(b) | Methods for subgroups and interactions | SDC p. 3 (Supplemental analyses); SDC p. 12 (Table S6). |
| 12(c) | Handling missing data | Manuscript pp. 4–5; SDC p. 7 (Table S1); SDC pp. 1–4 (Supplemental Methods). |
| 12(d) | Incomplete follow-up and censoring | Manuscript p. 5; SDC pp. 1–4 (Supplemental Methods); Manuscript p. 10. |
| 12(e) | Methods for sensitivity analyses | Manuscript p. 5; Table 3, Manuscript p. 17; SDC pp. 9–11 (Table S4). |
| 13(a) | Patient counts at successive stages | Manuscript pp. 6 and 8; SDC p. 5 (Figure S1); SDC pp. 11–12 (Table S5). |
| 13(b) | Reasons for exclusion or nonparticipation | Manuscript pp. 3–4; Manuscript p. 6; SDC p. 5 (Figure S1). |
| 13(c) | Consider a participant flow diagram | SDC p. 5 (Figure S1) |
| 14(a) | Clinical and demographic characteristics | Manuscript pp. 6–7; Table 1, Manuscript p. 15; Table 2, Manuscript p. 16. |
| 14(b) | Counts with missing values | SDC p. 7 (Table S1) |
| 14(c) | Summary of follow-up time | Manuscript p. 5; SDC p. 6 (Figure S3); SDC pp. 1–4 (Supplemental Methods). Follow-up horizons and at-risk counts are provided. |
| 15 | Outcome event counts or estimates over time | Manuscript pp. 6–8; Table 2, Manuscript p. 16; SDC pp. 8–9 (Table S3). |
| 16(a) | Main estimates and precision | Manuscript pp. 6–8; Table 2, Manuscript p. 16; Table 3, Manuscript p. 17. Estimates and intervals are reported for the descriptive comparisons. |

| Item | Reporting item | Location and reporting note |
|---|---|---|
| 16(b) | Boundaries for grouped quantitative variables | Manuscript p. 4; SDC pp. 1–4 (Supplemental Methods). |
| 16(c) | Absolute risks over a relevant horizon | Manuscript pp. 6–8; Table 2, Manuscript p. 16; SDC pp. 7–8 (Table S2). |
| 17 | Sensitivity and subgroup results | Manuscript pp. 7–8; Table 3, Manuscript p. 17; SDC p. 12 (Table S6); SDC pp. 9–11 (Table S4); SDC pp. 8–9 (Table S3). |
| 18 | Key findings in relation to objectives | Manuscript p. 8 |
| 19 | Limitations, bias and imprecision | Manuscript pp. 9–10 |
| 20 | Overall interpretation | Manuscript pp. 8–10 |
| 21 | Applicability outside the study population | Manuscript pp. 9–10. |
| 22 | Funding and role of funders | Manuscript p. 1 and p. 10. |

#### RECORD reporting checklist

Critically ill patients with cirrhosis outside the unified ACLF definition: a two-database cohort study

Manuscript page numbers refer to the medRxiv manuscript PDF. SDC pages refer to this supplement.

| Item | Reporting item | Location and reporting note |
|---|---|---|
| 1.1 | Type and names of routinely collected data | Manuscript p. 2 names both databases. Manuscript pp. 3–4 describe the data sources. |
| 1.2 | Setting and study years in the title or abstract | Manuscript pp. 3–4. Calendar years and setting are described in Methods. |
| 1.3 | Identification of cross-database linkage in abstract | Not applicable. The two databases were analyzed separately. |
| 6.1 | Population-selection algorithms and code lists | Manuscript pp. 3–4; SDC pp. 1–4 (Supplemental Methods); SDC p. 5 (Figure S1). |
| 6.2 | Evidence for eligibility algorithm validity | Manuscript pp. 3–4. The coding source is cited. Validation of the cohort identification algorithm in these databases was not performed. |
| 6.3 | Consider a study linkage diagram | SDC p. 5 (Figure S1). Internal table joins are described in SDC pp. 1–4 (Supplemental Methods); no cross-database linkage was performed. |
| 7.1 | Detailed codes or algorithms for study variables | Manuscript pp. 4–5; SDC pp. 1–4 (Supplemental Methods); SDC p. 7 (Table S1). |
| 12.1 | Scope of investigator access | Manuscript p. 6; Manuscript p. 10; SDC pp. 1–4 (Supplemental Methods). |
| 12.2 | Data cleaning procedures | Manuscript pp. 4–5; SDC pp. 1–4 (Supplemental Methods). |
| 12.3 | Linkage methods and quality if investigators linked databases | Manuscript p. 5; SDC pp. 1–4 (Supplemental Methods). MIMIC-IV includes source-provided death linkage. No linkage across the two study databases was performed. |
| 13.1 | Selection steps, including data availability | Manuscript pp. 6 and 8; SDC p. 5 (Figure S1); SDC pp. 11–12 (Table S5). |
| 19.1 | Limitations of routinely collected records | Manuscript pp. 9–10 |
| 22.1 | Availability of data, protocol and code | Manuscript p. 10. Cohort source, access restrictions and code repository are stated. |
